# Long COVID Risk and Pre-COVID Vaccination: An EHR-Based Cohort Study from the RECOVER Program

**DOI:** 10.1101/2022.10.06.22280795

**Authors:** M Daniel Brannock, Robert F Chew, Alexander J Preiss, Emily C Hadley, Julie A McMurry, Peter J Leese, Andrew T Girvin, Miles Crosskey, Andrea G Zhou, Richard A Moffitt, Michele Jonsson Funk, Emily R Pfaff, Melissa A Haendel, Christopher G Chute, the N3C and RECOVER Consortia

## Abstract

**Importance:** Characterizing the effect of vaccination on long COVID allows for better healthcare recommendations.

**Objective:** To determine if, and to what degree, vaccination prior to COVID-19 is associated with eventual long COVID onset, among those a documented COVID-19 infection.

**Design, Settings, and Participants:** Retrospective cohort study of adults with evidence of COVID-19 between August 1, 2021 and January 31, 2022 based on electronic health records from eleven healthcare institutions taking part in the NIH Researching COVID to Enhance Recovery (RECOVER) Initiative, a project of the National Covid Cohort Collaborative (N3C).

**Exposures:** Pre-COVID-19 receipt of a complete vaccine series versus no pre-COVID-19 vaccination.

**Main Outcomes and Measures:** Two approaches to the identification of long COVID were used. In the clinical diagnosis cohort (n=47,752), ICD-10 diagnosis codes or evidence of a healthcare encounter at a long COVID clinic were used. In the model-based cohort (n=199,498), a computable phenotype was used. The association between pre-COVID vaccination and long COVID was estimated using IPTW-adjusted logistic regression and Cox proportional hazards.

**Results:** In both cohorts, when adjusting for demographics and medical history, pre-COVID vaccination was associated with a reduced risk of long COVID (clinic-based cohort: HR, 0.66; 95% CI, 0.55-0.80; OR, 0.69; 95% CI, 0.59-0.82; model-based cohort: HR, 0.62; 95% CI, 0.56-0.69; OR, 0.70; 95% CI, 0.65-0.75).

**Conclusions and Relevance:** Long COVID has become a central concern for public health experts. Prior studies have considered the effect of vaccination on the prevalence of future long COVID symptoms, but ours is the first to thoroughly characterize the association between vaccination and clinically diagnosed or computationally derived long COVID. Our results bolster the growing consensus that vaccines retain protective effects against long COVID even in breakthrough infections.

**Key Points:** *Question:* Does vaccination prior to COVID-19 onset change the risk of long COVID diagnosis?

*Findings:* Four observational analyses of EHRs showed a statistically significant reduction in long COVID risk associated with pre-COVID vaccination (first cohort: HR, 0.66; 95% CI, 0.55-0.80; OR, 0.69; 95% CI, 0.59-0.82; second cohort: HR, 0.62; 95% CI, 0.56-0.69; OR, 0.70; 95% CI, 0.65-0.75).

*Meaning:* Vaccination prior to COVID onset has a protective association with long COVID even in the case of breakthrough infections.

## Introduction

The SARS-CoV-2 virus, and the COVID-19 pandemic it effected, hardly needs introducing more than two years after the WHO first announced evidence of human-to-human transmission in January of 2020.^1^ As of this writing, the WHO states there have been 594 million confirmed cases and more than 6 million deaths attributed to COVID-19 worldwide.^2^ Post-acute sequelae of SARS-CoV-2 infection (PASC) have been widely reported and can include any complication resulting from SARS-CoV-2 infection weeks after infection occurred.^3–5^ Long COVID is a single diagnosis that encapsulates the broad array of ever-shifting symptoms attributed to PASC.

Vaccines have been shown to be safe and effective at dramatically reducing the risk of severe COVID-19.^6,7^ Their impact on long COVID is less understood, with some studies indicating they have a significant protective effect^8–10^ while others reported mixed effects^11^ or even an anti-protective effect.^12^ While some have studied the impact of administering vaccines after the onset of PASC,^13–15^ we attempt to address ambiguity around the association between pre-COVID-19 vaccination and eventual long COVID diagnosis.

To our knowledge, we are the first to consider vaccination with long COVID directly, in the form of clinical diagnoses or a computable phenotype;^16^ previous studies have relied on the occurrence of one or two symptoms consistent with long or acute COVID. Ours is also the largest study to leverage time-to-event modeling or control for differences in the vaccinated and unvaccinated populations.

The National Institutes of Health (NIH) created the RECOVER initiative to address the uncertainty surrounding long COVID by coordinating research across hundreds of researchers and more than 30 institutions.^17^ The National COVID Cohort Collaborative (N3C),^18^ sponsored by NIH’s National Center for Advancing Translational Sciences, provides access to harmonized electronic health records (EHRs) through the N3C Data Enclave. More than 75 sites have contributed longitudinal data for over 15.5 million patients with a confirmed SARS-CoV-2 infection, COVID-19 symptoms, or their matched controls.

## Methods

### Base Population

This study is part of the NIH Researching COVID to Enhance Recovery (RECOVER) Initiative, which seeks to understand, treat, and prevent PASC. For more information on RECOVER, visit https://recovercovid.org. All analyses described here were performed within the secure N3C Data Enclave. N3C’s methods for patient identification, data acquisition, ingestion, data quality assessment, and harmonization have been described previously.^18,19^ The study population was drawn from 5,434,528 COVID-19-positive patients available in N3C. A COVID-19 index date (index) was defined as the earliest recorded indication of COVID-19 infection. Individuals who met the following inclusion criteria were eligible: (1) having an International Classification of Diseases-10-Clinical Modification (ICD-10) COVID-19 diagnosis code (U07.1) or a positive SARS-CoV-2 PCR or antigen test between August 1, 2021 and January 31, 2022; (2) having a recorded health care visit between 120 and 300 days after index; (3) having at least two recorded health care visits in the year prior to index; (4) being ≥18 years old at index; and (5) having either completed or not started a COVID-19 vaccine regimen at index.

One known limitation of EHR data is that only those healthcare encounters and services provided by the specific health system are available in the data.^20^ The proportion of patients with a recorded vaccination at a given health care site is driven by two factors: (1) the true rate of vaccination among the population served and (2) how consistently vaccines are captured by the site. Some sites report no vaccinations, while others sync vaccination records with their state’s vaccine registry. There is no explicit indicator of non-vaccination in the N3C Data Enclave, but sites with better vaccination coverage offer more confidence that patients with no recorded vaccine exposure are unvaccinated. We calculated vaccination coverage at each site as the ratio of two statistics: the observed proportion of patients with a vaccination record and an expected vaccination rate derived from CDC reporting^21^ for the population served. Sites with an observed proportion at least two-thirds of their expected vaccination rate were eligible for analysis, leaving 199,498 patients at eleven sites that met our inclusion criteria. A full breakdown of how many patients met our inclusion criteria is shown in Figure 1.

**Figure 1.**
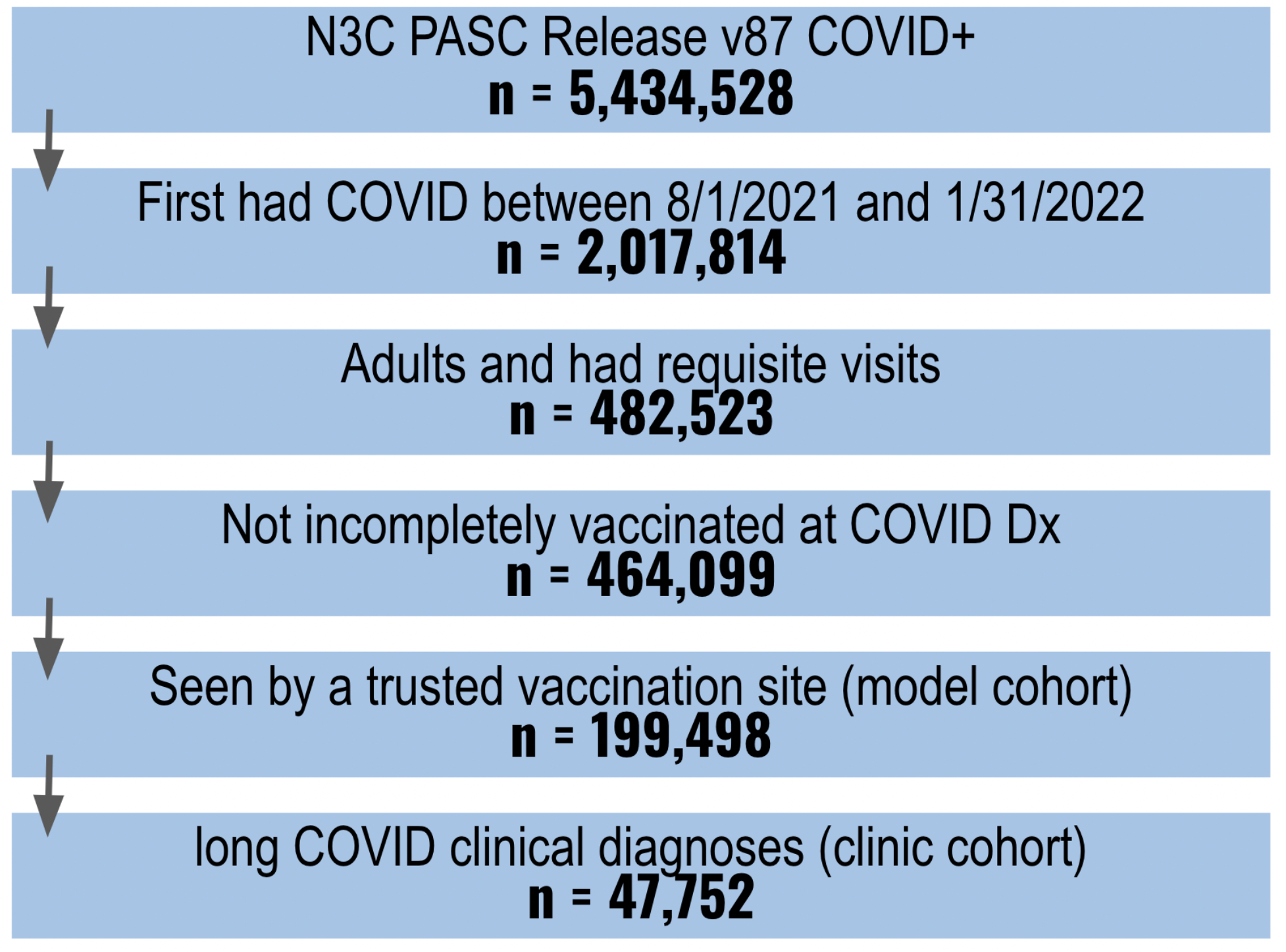
Cohort Definition Flowchart.

### Exposure Definition

Those who completed their vaccine regimen (2 mRNA or 1 viral vector vaccine) prior to index were considered vaccinated, while those with no recorded vaccines at index were considered unvaccinated. Partially vaccinated patients at index failed to meet the fifth inclusion criterion.

### Outcome Definitions

#### Clinical definition

We considered three clinical indicators of long COVID: (1) an ICD-10 code for post COVID-19 condition (U09.9), (2) an ICD-10 code for sequelae of other specific infectious and parasitic diseases (B94.8), or (3) a visit to a long COVID clinic. Prior to the introduction of U09.9 in October 2021, the CDC endorsed B94.8 to indicate long-term complications of SARS-CoV-2 infection. As with vaccination, not all sites report clinical indicators of long COVID. Six out of eleven sites, comprising 47,752 of 199,498 eligible patients, submitted clinical indicators of long COVID for at least 250 patients. We used patients from these six sites to form a clinic-based cohort of patients, whom we deemed eligible for receiving a clinical long COVID indicator.

Any long COVID clinical indicator was sufficient to label a patient as having had long COVID in the logistic regression. If patients had multiple encounters with a clinical indicator of long COVID, the earliest was used as the event date for purposes of the time-to-event analysis. Death and COVID-19 vaccination after COVID-19 onset were censoring events.

#### Model-based definition

Long COVID was classified in the model-based cohort using the long COVID cohort identification machine learning model (LC model) described in Pfaff et al, 2022;^16^ the model was retrained with U09.9 diagnoses as the target event and without vaccination status as an input.

The model calculates a long COVID likelihood score (range 0 to 1) for each patient beginning 100 days after index using only conditions and drugs observed as of that day. New scores are generated in 30-day intervals until 300 days after index or June 1, 2022, whichever comes first. Patients scoring above 0.9 in any interval were labeled as having long COVID. A threshold of 0.9 was chosen as it resulted in a similar prevalence of long COVID across the model-based and clinic-based cohorts. The earliest interval receiving a score above 0.9 was assigned as the event date for purposes of the time-to-event analysis. As in the clinic-based definition, death and COVID-19 vaccination were censoring events.

Any patient meeting our inclusion criteria from any of the eleven sites was eligible for a model-derived indicator of long COVID and was included in the model-based cohort. Therefore, all patients in the clinic-based cohort are also included in the model-based cohort, where they can (and sometimes do) have a different assigned long COVID outcome. This is not unexpected— the LC model was trained using U09.9 as the target, while we include U09.9, B94.8, and long COVID clinic visits as valid clinical diagnoses. Both labels are rare and imperfect; we do not expect one indication to guarantee the other.

### Statistical Analysis

Two analyses were carried out to estimate the association between vaccination and long COVID: (1) logistic regression to calculate an overall association while controlling for patient characteristics, and (2) Cox proportional hazards to incorporate potential differences in the time-to-event for long COVID. We do not consider either analysis as primary, as each has weaknesses addressed by the other. Proportional hazards requires a date for long COVID diagnosis and for hazard functions to be proportional over time. Both are difficult to fully validate, and logistic regression requires neither. Logistic regression fails to consider varying times-to-event and vaccinations after COVID-19, which are accounted for in proportional hazards. We present the results of both analyses as a test of the robustness of the association.

Inverse probability of treatment weighting (IPTW) was applied to both logistic regression and proportional hazards to control for differences in patient characteristics across the vaccinated and unvaccinated groups. Logistic regression was used to estimate the propensity score based on demographics, medical history, social determinants of health, and spatial and temporal variables. Our selection of covariates was informed by the literature on important indicators of long COVID and is shown in eTables 1 and 2.^16,22,23^ Covariate balance before and after weighting was evaluated with standardized mean differences. Covariates with a standardized mean difference less than 0.1 were considered well-balanced. Stabilized treatment weights were calculated as outlined in Robins et al (2000).^24^ Standard errors in the IPTW-adjusted models were calculated from 200 bootstrapped iterations based on the standard deviation of the estimates.^25^ Unadjusted associations were also calculated and reported.

For logistic regression models, studentized residuals, leverage scores, Cook’s distances, and DFBETAS were examined to identify influential observations. For proportional hazards models, the Lifelines package’s *CoxPHFitter*.*check_assumptions* method was used to test the assumption that each covariate’s effect on the hazard rate is constant over time.^26,27^ Interactions with time were added to the model for covariates which did not meet the proportional hazards assumption.

#### Sensitivity analyses

Sensitivity of the IPTW-adjusted and unadjusted vaccination status coefficients in the logistic regression and proportional hazards models were tested across three dimensions: (1) LC model threshold (0.3 to 0.95), (2) with or without independent features in addition to vaccination, and (3) including or not including post-index vaccinations as a censoring event. The first sensitivity dimension was not relevant in the clinic-based cohort and the third was not relevant for logistic regression analyses.

All analyses were conducted using Python (version 3.6.10) with the Statsmodels (0.12.2) and Lifelines (0.26.4) packages. Study design elements, methods, and results were reported consistent with STROBE guidelines.^28^

## Results

A summary of patient characteristics for both cohorts is shown in Table 1. The IPTW-adjusted logistic regression and proportional hazards models showed strong, protective associations in both cohorts (Table 2). The full tables of coefficients are provided as eTables 3–6 in the online supplement. There was not a clear association between vaccination status and long COVID in the unadjusted model-based analysis, though an association could still be observed in the clinic-based cohort (Table 2). The IPTW-adjusted Kaplan-Meier curves for the model-based and clinic-based cohorts are shown in Figure 2.

**Table 1.** Baseline Patient Characteristics.

| Variable | Model-Based Cohort |  |  | Clinic-Based Cohort |  |  |
| --- | --- | --- | --- | --- | --- | --- |
|  | Overall<br>(N=199498) | Fully<br>Vaccinated<br>(N=87099) | Unvaccinated<br>(N=112399) | Overall<br>(N=47752) | Fully<br>Vaccinated<br>(N=26567) | Unvaccinated<br>(N=21185) |
| Mean Age | 47.22 (100.0) | 52.46 (100.0) | 43.16 (100.0) | 48.17 (100.0) | 50.97 (100.0) | 44.65 (100.0) |
| Gender |  |  |  |  |  |  |
| Female | 128920 (64.6) | 56561 (64.9) | 72359 (64.4) | 31051 (65.0) | 17499 (65.9) | 13552 (64.0) |
| Male | 70578 (35.4) | 30538 (35.1) | 40040 (35.6) | 16701 (35.0) | 9068 (34.1) | 7633 (36.0) |
| Age at COVID Index Date |  |  |  |  |  |  |
| 18-24 | 20701 (10.4) | 4922 (5.7) | 15779 (14.0) | 4522 (9.5) | 1753 (6.6) | 2769 (13.1) |
| 25-34 | 36729 (18.4) | 11382 (13.1) | 25347 (22.6) | 8557 (17.9) | 4078 (15.3) | 4479 (21.1) |
| 35-49 | 53883 (27.0) | 21646 (24.9) | 32237 (28.7) | 12445 (26.1) | 6748 (25.4) | 5697 (26.9) |
| 50-64 | 50887 (25.5) | 25332 (29.1) | 25555 (22.7) | 12356 (25.9) | 7235 (27.2) | 5121 (24.2) |
| 65+ | 37298 (18.7) | 23817 (27.3) | 13481 (12.0) | 9872 (20.7) | 6753 (25.4) | 3119 (14.7) |
| Race / Ethnicity |  |  |  |  |  |  |
| Asian Non-Hispanic | 3542 (1.8) | 2625 (3.0) | 917 (0.8) | 861 (1.8) | 670 (2.5) | 191 (0.9) |
| Black or African American Non-Hispanic | 26588 (13.3) | 10330 (11.9) | 16258 (14.5) | 10588 (22.2) | 5355 (20.2) | 5233 (24.7) |
| Hispanic or Latino Any Race | 18870 (9.5) | 9876 (11.3) | 8994 (8.0) | 3089 (6.5) | 1642 (6.2) | 1447 (6.8) |
| NHOPI Non-Hispanic | 272 (0.1) | 156 (0.2) | 116 (0.1) | 75 (0.2) | 38 (0.1) | 37 (0.2) |
| Other Non-Hispanic | 4236 (2.1) | 1477 (1.7) | 2759 (2.5) | 1391 (2.9) | 460 (1.7) | 931 (4.4) |
| Unknown | 2693 (1.3) | 1490 (1.7) | 1203 (1.1) | 1172 (2.5) | 626 (2.4) | 546 (2.6) |
| White Non-Hispanic | 143297 (71.8) | 61145 (70.2) | 82152 (73.1) | 30576 (64.0) | 17776 (66.9) | 12800 (60.4) |
| Data Partner |  |  |  |  |  |  |
| Partner A | 9526 (4.8) | 5853 (6.7) | 3673 (3.3) | 9367 (19.6) | 5776 (21.7) | 3591 (17.0) |
| Partner B | 1746 (0.9) | 1162 (1.3) | 584 (0.5) |  |  |  |
| Partner C | 3449 (1.7) | 2114 (2.4) | 1335 (1.2) | 3414 (7.1) | 2093 (7.9) | 1321 (6.2) |
| Partner D | 1176 (0.6) | 754 (0.9) | 422 (0.4) | 1170 (2.5) | 752 (2.8) | 418 (2.0) |
| Partner E | 2734 (1.4) | 2092 (2.4) | 642 (0.6) |  |  |  |
| Partner F | 27322 (13.7) | 14857 (17.1) | 12465 (11.1) |  |  |  |
| Partner G | 6095 (3.1) | 3984 (4.6) | 2111 (1.9) | 6008 (12.6) | 3926 (14.8) | 2082 (9.8) |
| Partner H | 2136 (1.1) | 997 (1.1) | 1139 (1.0) |  |  |  |
| Partner I | 2281 (1.1) | 1460 (1.7) | 821 (0.7) | 2215 (4.6) | 1424 (5.4) | 791 (3.7) |
| Partner J | 25946 (13.0) | 12774 (14.7) | 13172 (11.7) | 25578 (53.6) | 12596 (47.4) | 12982 (61.3) |
| Partner K | 117087 (58.7) | 41052 (47.1) | 76035 (67.6) |  |  |  |
| COVID Month |  |  |  |  |  |  |
| August 2021 | 48333 (24.2) | 15273 (17.5) | 33060 (29.4) | 7440 (15.6) | 2893 (10.9) | 4547 (21.5) |
| September 2021 | 45601 (22.9) | 16116 (18.5) | 29485 (26.2) | 7314 (15.3) | 3176 (12.0) | 4138 (19.5) |
| October 2021 | 22949 (11.5) | 9382 (10.8) | 13567 (12.1) | 3482 (7.3) | 1705 (6.4) | 1777 (8.4) |
| November 2021 | 24074 (12.1) | 10202 (11.7) | 13872 (12.3) | 2942 (6.2) | 1505 (5.7) | 1437 (6.8) |
| December 2021 | 23321 (11.7) | 12150 (13.9) | 11171 (9.9) | 7525 (15.8) | 4507 (17.0) | 3018 (14.2) |
| January 2022 | 35220 (17.7) | 23976 (27.5) | 11244 (10.0) | 19049 (39.9) | 12781 (48.1) | 6268 (29.6) |
| Health Status |  |  |  |  |  |  |
| Immunocompromised | 2153 (1.1) | 1544 (1.8) | 609 (0.5) | 1300 (2.7) | 1009 (3.8) | 291 (1.4) |
| Diabetes (Complicated) | 19805 (9.9) | 11412 (13.1) | 8393 (7.5) | 6715 (14.1) | 4293 (16.2) | 2422 (11.4) |
| Diabetes (Uncomplicated) | 30550 (15.3) | 17044 (19.6) | 13506 (12.0) | 9833 (20.6) | 6106 (23.0) | 3727 (17.6) |
| Kidney Disease | 13211 (6.6) | 8028 (9.2) | 5183 (4.6) | 5259 (11.0) | 3463 (13.0) | 1796 (8.5) |
| Acute Kidney Injury | 7614 (3.8) | 4281 (4.9) | 3333 (3.0) | 3562 (7.5) | 2183 (8.2) | 1379 (6.5) |
| Chronic Lung Disease | 26652 (13.4) | 13964 (16.0) | 12688 (11.3) | 10261 (21.5) | 6089 (22.9) | 4172 (19.7) |
| Tobacco Smoker | 8375 (4.2) | 3205 (3.7) | 5170 (4.6) | 5065 (10.6) | 2061 (7.8) | 3004 (14.2) |
| Heart Failure | 8281 (4.2) | 4925 (5.7) | 3356 (3.0) | 3442 (7.2) | 2199 (8.3) | 1243 (5.9) |
| Myocardial Infarction | 4950 (2.5) | 2726 (3.1) | 2224 (2.0) | 2225 (4.7) | 1314 (4.9) | 911 (4.3) |
| Congestive Heart Failure | 6450 (3.2) | 3932 (4.5) | 2518 (2.2) | 2813 (5.9) | 1827 (6.9) | 986 (4.7) |

**Table 2a.** Long COVID by Vaccination Status: Measures of Association.

|  |  | Logistic Regression<br>OR<sup>a</sup> (95% CI) | Proportional Hazards<br>HR<sup>b</sup> (95% CI) |
| --- | --- | --- | --- |
| IPTW-Adjusted | Model-Based Cohort | 0.70 (0.65, 0.75) | 0.62 (0.56, 0.69) |
|  | Clinic-Based Cohort | 0.69 (0.59, 0.82) | 0.66 (0.55, 0.80) |
| Unadjusted | Model-Based Cohort | 1.04 (0.97, 1.11) | 0.99 (0.90, 1.08) |
|  | Clinic-Based Cohort | 0.79 (0.68, 0.92) | 0.80 (0.67, 0.95) |

<sup>a</sup>OR: Odds Ratio, <sup>b</sup>HR: Hazard Ratio

**Table 2b.** Long COVID by Vaccination Status: Unadjusted Counts.

|  | Model-Based Cohort |  |  | Clinic-Based Cohort |  |  |
| --- | --- | --- | --- | --- | --- | --- |
|  | Overall | With Long COVID | Without Long COVID | Overall | With Long COVID | Without Long COVID |
| Fully Vaccinated | 87,099 (100%) | 1,516 (1.7%) | 85,583 (98.3%) | 26,567 (100%) | 352 (1.3%) | 26,215 (98.7%) |
| Unvaccinated | 112,399 (100%) | 1,889 (1.7%) | 110,510 (98.3%) | 21,185 (100%) | 354 (1.7%) | 20,831 (98.3%) |

**Figure 2.**
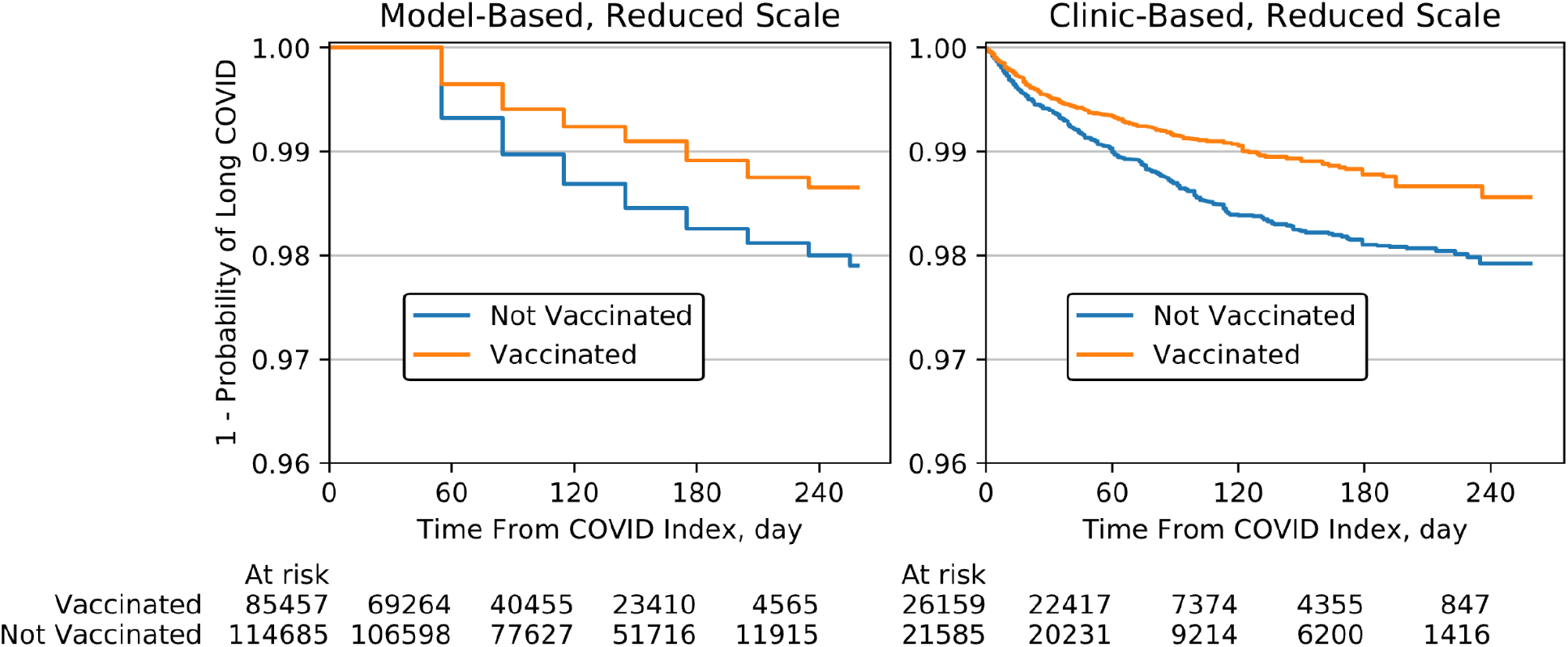
IPTW-Adjusted Kaplan-Meier Curves. Long COVID events can only be observed in the model-based cohort in 30-day increments, resulting in the observed stair-step structure. A reduced scale is used to highlight the differentiation between the vaccinated and unvaccinated curves.

Key results of the sensitivity analysis are summarized in Figure 3. The association between vaccination and long COVID was robust to excluding either IPTW adjustment or non-vaccination covariates, but not both. In the proportional hazards models, the association was robust to not censoring post-COVID-19 vaccination events (uncensored points are not pictured in Figure 3 as they closely overlap the censored points). In the model-based cohort, the association was not robust to the LC model threshold, with lower thresholds resulting in a progressively weaker protective association.

**Figure 3.**
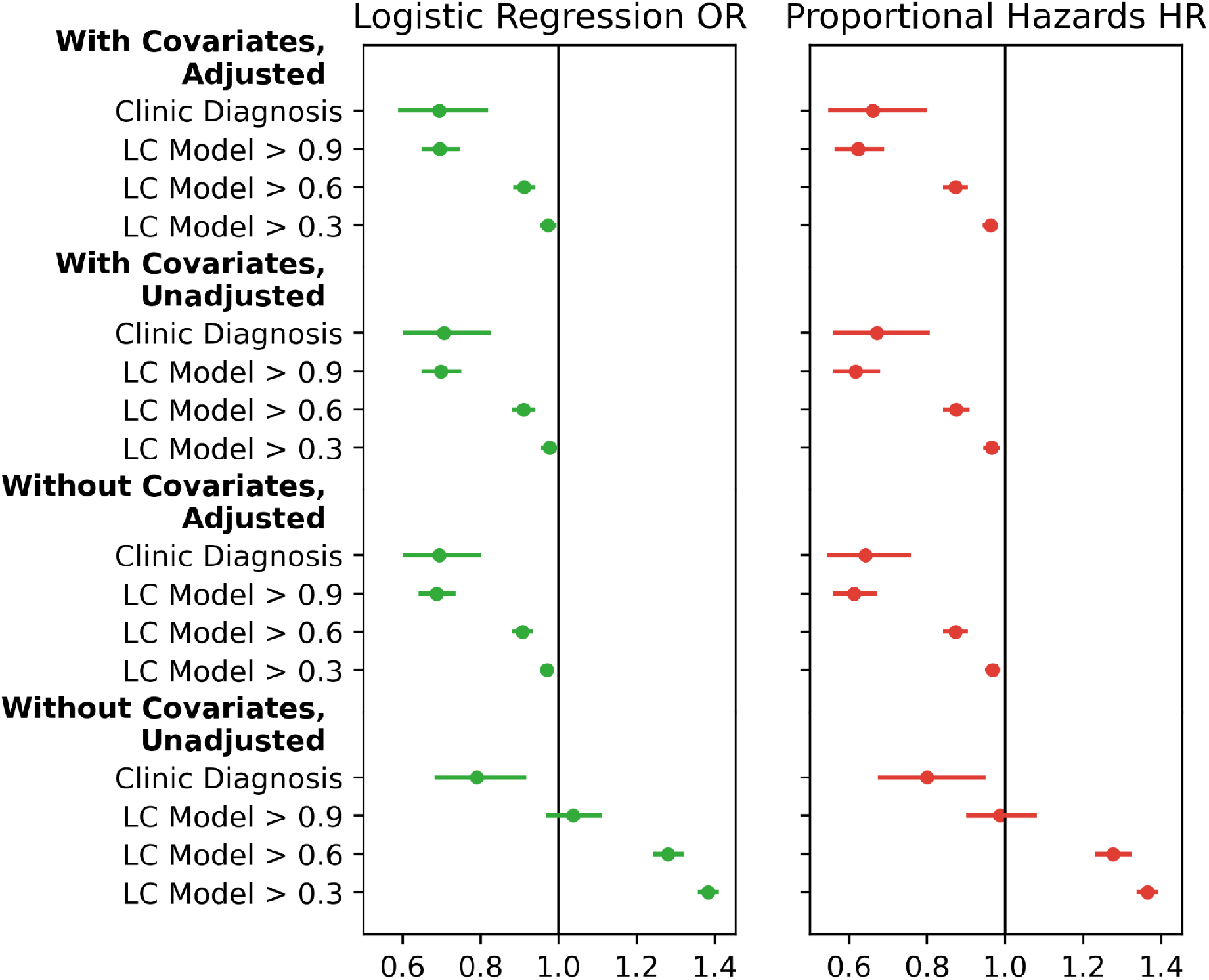
Sensitivity analysis vaccination coefficients for logistic regression (LR) and proportional hazards (PH). Odds ratios (OR) are shown for LR, hazard ratios (HR) are shown for PH.

After IPTW-adjustment, all covariates were well-balanced (eFigures 1 and 2). Logistic regression diagnostics did not indicate any overly influential observations. Observations with large residuals tended to have low leverage and vice versa. In the model-based cohort, the greatest Cook’s distance was < 0.01 and the greatest absolute DFBETA for vaccination status was 0.04. In the clinic-based cohort, the greatest Cook’s distance was 0.01 and the greatest absolute DFBETA for vaccination status was 0.08. In the model-based cohort, seven patients had stabilized inverse probability of treatment weights above 20 (max of 33); excluding these patients did not impact vaccination coefficients at the precision reported here. The maximum weight in the clinic-based cohort was nine.

## Discussion

Our four analyses yielded consistent results. We see protective associations of vaccination with long COVID onset in both logistic and time-to-event models, and in both clinic-based and model-based cohorts. While these findings are similar to those of other large observational studies,^8–10^ previous sources have only looked for evidence of COVID-associated symptoms as evidence of long COVID. A major finding of our analysis is that the protective association remains consistent in results where a clinical diagnosis is required, and among those who contracted COVID-19 in a later period that includes Omicron infections. The use of the LC model allowed us to expand our sample from six to eleven sites and 47,752 to 199,498 COVID-positive patients, across which we confirmed consistent results.

Interestingly, the protective association of vaccination with long COVID is weaker or reversed in the unadjusted coefficients and cross tabulations (Table 2, Figure 3). Several features that are associated with a higher likelihood of long COVID (coefficients in eTables 3–6) are also associated with a higher likelihood of vaccination (coefficients in eTables 1–2). The most significant is age: eTable 7 shows how older adults are both more likely to be vaccinated and more likely to contract long COVID in comparison to younger adults. Failing to account for the substantial differences between individuals who were and were not vaccinated prior to COVID-19 could lead one to conclude that vaccination is harmful.

The sensitivity analysis presents other instructive complexities. Reducing the LC model threshold lowers the amount of evidence required to denote someone as having long COVID; it also moderates the protective association of vaccination with long COVID (key results in Figure 3, full range of thresholds in eFigure 3). While we’d expect that including healthy adults in the long COVID population would dilute the observed protective association, individuals with a LC model score between 0.6 and 0.9 are not entirely healthy—they have some evidence of long COVID. If high-confidence and clinically diagnosed long COVID cases are more severe than cases with fewer recorded symptoms, it could suggest that vaccination is most strongly associated with a reduced risk of severe long COVID. More work is needed to validate that conclusion.

Healthcare utilization is one of the most important features in the LC model.^16^ If fully vaccinated patients are more likely to utilize the healthcare system, the LC model’s marginal predictions may be assigning more fully vaccinated individuals to long COVID because they are more likely to interact with the healthcare system, depressing the observed benefit of vaccination. A known challenge of analyzing EHR data is that they tend to provide more information on individuals who regularly utilize healthcare systems,^29^ though we attempt to control for this by requiring multiple recorded encounters outside of COVID-19 for inclusion in the study.

It is well-documented that vaccination reduces the risk of developing COVID-19,^6,7^ offering one mechanism for preventing Long COVID. However, there is evidence that widely circulated vaccines are less effective against now-dominant Omicron than earlier SARS-CoV-2 variants,^30– 32^ increasing interest in whether or not vaccination reduces the risk of long COVID in breakthrough infections. That is the aim of this study, in which all eligible patients had a COVID-19 diagnosis. As a result, the stated association between vaccination and long COVID will be an underestimate of the effective association in the general population due to the primary prevention of COVID-19 in the first place.

IPTW is often used to estimate causal effects from observational data and is employed here to provide more robust associations. However, we do not interpret these results as causal effects. This is for two reasons: (1) we are unwilling to assume that there are no unmeasured confounders in our treatment model and (2) our causal model includes several latent variables, which obstruct the estimation of treatment effects through covariate adjustment. We explore each reason in the eDiscussion of the online supplement and provide a directed acyclic graph of confounders in eFigure 4.

## Limitations

Our study is limited by its reliance on EHRs and other factors. Those who choose to not seek healthcare, or are unable to do so, are not represented in EHRs. Even among available patients, our sample is biased towards high utilizers and those with hospitalizations. We are forced to assume that those without a recorded condition or symptom do not exhibit it, including potentially unrecorded reinfections of COVID-19. We attempt to mitigate this limitation with respect to vaccination by carefully selecting healthcare sites with reasonably high reported vaccination rates, but some vaccinations remain unreported, likely resulting in a conservative estimate.

We did not distinguish between vaccine types, though previous studies and initial tabulations failed to detect differences in their effectiveness in preventing long COVID.^9,10^

The ICD10 code for long COVID, U09.9, was not implemented until October 2021, and its full adoption was not immediate. The previously recommended ICD10 code, B94.8, is more general and is used to diagnose long-term complications from any viral infection. We accepted B94.8 as a long COVID diagnosis because use of the code in our data by mid-2021 was 40 times higher than its baseline use in 2018 and 2019.

Finally, the confidence intervals around the LC model-based risk estimates are likely to narrow as there remains residual misclassification of Long COVID outcomes in that cohort not factored into the confidence interval boundaries.

## Conclusions

Vaccination is a proven tool in combating onset of COVID-19. We show that benefits of vaccination persist in breakthrough infections through a moderate but consistent protective association against clinically diagnosed long COVID.

## Data Availability

All data used in this study is available through the N3C Enclave to approved users. See https://covid.cd2h.org/for-researchers for instructions on how to access the data.

## Acknowledgements

Authorship was determined using ICMJE recommendations. The content is solely the responsibility of the authors and does not necessarily represent the official views of the National Institutes of Health or N3C or RECOVER.

This study is part of the NIH Researching COVID to Enhance Recovery (RECOVER) Initiative, which seeks to understand, treat, and prevent the post-acute sequelae of SARS-CoV-2 infection (PASC). For more information on RECOVER, visit https://recovercovid.org/. This research was funded by the National Institutes of Health (NIH) Agreement OTA OT2HL161847 as part of the Researching COVID to Enhance Recovery (RECOVER) research program.

We would like to thank the National Community Engagement Group (NCEG), all patient, caregiver, and community Representatives, and all the participants enrolled in the RECOVER Initiative.

## N3C Attribution

The analyses described in this publication were conducted with data or tools accessed through the NCATS N3C Data Enclave covid.cd2h.org/enclave and supported by CD2H - The National COVID Cohort Collaborative (N3C) IDeA CTR Collaboration 3U24TR002306-04S2 NCATS U24 TR002306. This research was possible because of the patients whose information is included within the data from participating organizations (covid.cd2h.org/dtas) and the organizations and scientists (covid.cd2h.org/duas) who have contributed to the on-going development of this community resource (cite this https://doi.org/10.1093/jamia/ocaa196).

## IRB

The N3C data transfer to NCATS is performed under a Johns Hopkins University Reliance Protocol # IRB00249128 or individual site agreements with NIH. The N3C Data Enclave is managed under the authority of the NIH; information can be found at https://ncats.nih.gov/n3c/resources.

## We gratefully acknowledge the following core contributors to N3C

Adam B. Wilcox, Adam M. Lee, Alexis Graves, Alfred (Jerrod) Anzalone, Amin Manna, Amit Saha, Amy Olex, Andrea Zhou, Andrew E. Williams, Andrew Southerland, Andrew T. Girvin, Anita Walden, Anjali A. Sharathkumar, Benjamin Amor, Benjamin Bates, Brian Hendricks, Brijesh Patel, Caleb Alexander, Carolyn Bramante, Cavin Ward-Caviness, Charisse Madlock-Brown, Christine Suver, Christopher Chute, Christopher Dillon, Chunlei Wu, Clare Schmitt, Cliff Takemoto, Dan Housman, Davera Gabriel, David A. Eichmann, Diego Mazzotti, Don Brown, Eilis Boudreau, Elaine Hill, Elizabeth Zampino, Emily Carlson Marti, Emily R. Pfaff, Evan French, Farrukh M Koraishy, Federico Mariona, Fred Prior, George Sokos, Greg Martin, Harold Lehmann, Heidi Spratt, Hemalkumar Mehta, Hongfang Liu, Hythem Sidky, J.W. Awori Hayanga, Jami Pincavitch, Jaylyn Clark, Jeremy Richard Harper, Jessica Islam, Jin Ge, Joel Gagnier, Joel H. Saltz, Joel Saltz, Johanna Loomba, John Buse, Jomol Mathew, Joni L. Rutter, Julie A. McMurry, Justin Guinney, Justin Starren, Karen Crowley, Katie Rebecca Bradwell, Kellie M. Walters, Ken Wilkins, Kenneth R. Gersing, Kenrick Dwain Cato, Kimberly Murray, Kristin Kostka, Lavance Northington, Lee Allan Pyles, Leonie Misquitta, Lesley Cottrell, Lili Portilla, Mariam Deacy, Mark M. Bissell, Marshall Clark, Mary Emmett, Mary Morrison Saltz, Matvey B. Palchuk, Melissa A. Haendel, Meredith Adams, Meredith Temple-O’Connor, Michael G. Kurilla, Michele Morris, Nabeel Qureshi, Nasia Safdar, Nicole Garbarini, Noha Sharafeldin, Ofer Sadan, Patricia A. Francis, Penny Wung Burgoon, Peter Robinson, Philip R.O. Payne, Rafael Fuentes, Randeep Jawa, Rebecca Erwin-Cohen, Rena Patel, Richard A. Moffitt, Richard L. Zhu, Rishi Kamaleswaran, Robert Hurley, Robert T. Miller, Saiju Pyarajan, Sam G. Michael, Samuel Bozzette, Sandeep Mallipattu, Satyanarayana Vedula, Scott Chapman, Shawn T. O’Neil, Soko Setoguchi, Stephanie S. Hong, Steve Johnson, Tellen D. Bennett, Tiffany Callahan, Umit Topaloglu, Usman Sheikh, Valery Gordon, Vignesh Subbian, Warren A. Kibbe, Wenndy Hernandez, Will Beasley, Will Cooper, William Hillegass, Xiaohan Tanner Zhang. Details of contributions available at covid.cd2h.org/core-contributors

## Data Partners with Released Data

The following institutions whose data is released or pending:

Available: Advocate Health Care Network — UL1TR002389: The Institute for Translational Medicine (ITM) • Boston University Medical Campus — UL1TR001430: Boston University Clinical and Translational Science Institute • Brown University — U54GM115677: Advance Clinical Translational Research (Advance-CTR) • Carilion Clinic — UL1TR003015: iTHRIV Integrated Translational health Research Institute of Virginia • Charleston Area Medical Center — U54GM104942: West Virginia Clinical and Translational Science Institute (WVCTSI) • Children’s Hospital Colorado — UL1TR002535: Colorado Clinical and Translational Sciences Institute • Columbia University Irving Medical Center — UL1TR001873: Irving Institute for Clinical and Translational Research • Duke University — UL1TR002553: Duke Clinical and Translational Science Institute • George Washington Children’s Research Institute — UL1TR001876: Clinical and Translational Science Institute at Children’s National (CTSA-CN) • George Washington University — UL1TR001876: Clinical and Translational Science Institute at Children’s National (CTSA-CN) • Indiana University School of Medicine — UL1TR002529: Indiana Clinical and Translational Science Institute • Johns Hopkins University — UL1TR003098: Johns Hopkins Institute for Clinical and Translational Research • Loyola Medicine — Loyola University Medical Center • Loyola University Medical Center — UL1TR002389: The Institute for Translational Medicine (ITM) • Maine Medical Center — U54GM115516: Northern New England Clinical & Translational Research (NNE-CTR) Network • Massachusetts General Brigham — UL1TR002541: Harvard Catalyst • Mayo Clinic Rochester — UL1TR002377: Mayo Clinic Center for Clinical and Translational Science (CCaTS) • Medical University of South Carolina — UL1TR001450: South Carolina Clinical & Translational Research Institute (SCTR) • Montefiore Medical Center — UL1TR002556: Institute for Clinical and Translational Research at Einstein and Montefiore • Nemours — U54GM104941: Delaware CTR ACCEL Program • NorthShore University HealthSystem — UL1TR002389: The Institute for Translational Medicine (ITM) • Northwestern University at Chicago — UL1TR001422: Northwestern University Clinical and Translational Science Institute (NUCATS) • OCHIN — INV-018455: Bill and Melinda Gates Foundation grant to Sage Bionetworks • Oregon Health & Science University — UL1TR002369: Oregon Clinical and Translational Research Institute • Penn State Health Milton S. Hershey Medical Center — UL1TR002014: Penn State Clinical and Translational Science Institute • Rush University Medical Center — UL1TR002389: The Institute for Translational Medicine (ITM) • Rutgers, The State University of New Jersey — UL1TR003017: New Jersey Alliance for Clinical and Translational Science • Stony Brook University — U24TR002306 • The Ohio State University — UL1TR002733: Center for Clinical and Translational Science • The State University of New York at Buffalo — UL1TR001412: Clinical and Translational Science Institute • The University of Chicago — UL1TR002389: The Institute for Translational Medicine (ITM) • The University of Iowa — UL1TR002537: Institute for Clinical and Translational Science • The University of Miami Leonard M. Miller School of Medicine — UL1TR002736: University of Miami Clinical and Translational Science Institute • The University of Michigan at Ann Arbor — UL1TR002240: Michigan Institute for Clinical and Health Research • The University of Texas Health Science Center at Houston — UL1TR003167: Center for Clinical and Translational Sciences (CCTS) • The University of Texas Medical Branch at Galveston — UL1TR001439: The Institute for Translational Sciences • The University of Utah — UL1TR002538: Uhealth Center for Clinical and Translational Science • Tufts Medical Center — UL1TR002544: Tufts Clinical and Translational Science Institute • Tulane University — UL1TR003096: Center for Clinical and Translational Science • University Medical Center New Orleans — U54GM104940: Louisiana Clinical and Translational Science (LA CaTS) Center • University of Alabama at Birmingham — UL1TR003096: Center for Clinical and Translational Science • University of Arkansas for Medical Sciences — UL1TR003107: UAMS Translational Research Institute • University of Cincinnati — UL1TR001425: Center for Clinical and Translational Science and Training • University of Colorado Denver, Anschutz Medical Campus — UL1TR002535: Colorado Clinical and Translational Sciences Institute • University of Illinois at Chicago — UL1TR002003: UIC Center for Clinical and Translational Science • University of Kansas Medical Center — UL1TR002366: Frontiers: University of Kansas Clinical and Translational Science Institute • University of Kentucky — UL1TR001998: UK Center for Clinical and Translational Science • University of Massachusetts Medical School Worcester — UL1TR001453: The UMass Center for Clinical and Translational Science (UMCCTS) • University of Minnesota — UL1TR002494: Clinical and Translational Science Institute • University of Mississippi Medical Center — U54GM115428: Mississippi Center for Clinical and Translational Research (CCTR) • University of Nebraska Medical Center — U54GM115458: Great Plains IDeA-Clinical & Translational Research • University of North Carolina at Chapel Hill — UL1TR002489: North Carolina Translational and Clinical Science Institute • University of Oklahoma Health Sciences Center — U54GM104938: Oklahoma Clinical and Translational Science Institute (OCTSI) • University of Rochester — UL1TR002001: UR Clinical & Translational Science Institute • University of Southern California — UL1TR001855: The Southern California Clinical and Translational Science Institute (SC CTSI) • University of Vermont — U54GM115516: Northern New England Clinical & Translational Research (NNE-CTR) Network • University of Virginia — UL1TR003015: iTHRIV Integrated Translational health Research Institute of Virginia • University of Washington — UL1TR002319: Institute of Translational Health Sciences • University of Wisconsin-Madison — UL1TR002373: UW Institute for Clinical and Translational Research • Vanderbilt University Medical Center — UL1TR002243: Vanderbilt Institute for Clinical and Translational Research • Virginia Commonwealth University — UL1TR002649: C. Kenneth and Dianne Wright Center for Clinical and Translational Research • Wake Forest University Health Sciences — UL1TR001420: Wake Forest Clinical and Translational Science Institute • Washington University in St. Louis — UL1TR002345: Institute of Clinical and Translational Sciences • Weill Medical College of Cornell University — UL1TR002384: Weill Cornell Medicine Clinical and Translational Science Center • West Virginia University — U54GM104942: West Virginia Clinical and Translational Science Institute (WVCTSI)

### Submitted

Icahn School of Medicine at Mount Sinai — UL1TR001433: ConduITS Institute for Translational Sciences • The University of Texas Health Science Center at Tyler — UL1TR003167: Center for Clinical and Translational Sciences (CCTS) • University of California, Davis — UL1TR001860: UCDavis Health Clinical and Translational Science Center • University of California, Irvine — UL1TR001414: The UC Irvine Institute for Clinical and Translational Science (ICTS) • University of California, Los Angeles — UL1TR001881: UCLA Clinical Translational Science Institute • University of California, San Diego — UL1TR001442: Altman Clinical and Translational Research Institute • University of California, San Francisco — UL1TR001872: UCSF Clinical and Translational Science Institute Pending: Arkansas Children’s Hospital — UL1TR003107: UAMS Translational Research Institute • Baylor College of Medicine — None (Voluntary) • Children’s Hospital of Philadelphia — UL1TR001878: Institute for Translational Medicine and Therapeutics • Cincinnati Children’s Hospital Medical Center — UL1TR001425: Center for Clinical and Translational Science and Training • Emory University — UL1TR002378: Georgia Clinical and Translational Science Alliance • HonorHealth — None (Voluntary) • Loyola University Chicago — UL1TR002389: The Institute for Translational Medicine (ITM) • Medical College of Wisconsin — UL1TR001436: Clinical and Translational Science Institute of Southeast Wisconsin • MedStar Health Research Institute — UL1TR001409: The Georgetown-Howard Universities Center for Clinical and Translational Science (GHUCCTS) • MetroHealth — None (Voluntary) • Montana State University — U54GM115371: American Indian/Alaska Native CTR • NYU Langone Medical Center — UL1TR001445: Langone Health’s Clinical and Translational Science Institute • Ochsner Medical Center — U54GM104940: Louisiana Clinical and Translational Science (LA CaTS) Center • Regenstrief Institute — UL1TR002529: Indiana Clinical and Translational Science Institute • Sanford Research — None (Voluntary) • Stanford University — UL1TR003142: Spectrum: The Stanford Center for Clinical and Translational Research and Education • The Rockefeller University — UL1TR001866: Center for Clinical and Translational Science • The Scripps Research Institute — UL1TR002550: Scripps Research Translational Institute • University of Florida — UL1TR001427: UF Clinical and Translational Science Institute • University of New Mexico Health Sciences Center — UL1TR001449: University of New Mexico Clinical and Translational Science Center • University of Texas Health Science Center at San Antonio — UL1TR002645: Institute for Integration of Medicine and Science • Yale New Haven Hospital — UL1TR001863: Yale Center for Clinical Investigation

