## Supplementary material for "Long COVID Risk and Pre-COVID Vaccination: An EHR-Based Cohort Study from the RECOVER Program": Online Supplement

**eTable 1. Model-Based IPTW Model Coefficients**

| Covariate | Coefficient | Standard Error | 95% CI Low | 95% CI High |
|---|---|---|---|---|
| Age: 18-24 | -0.79 | 0.02 | -0.83 | -0.75 |
| Age: 25-34 | -0.43 | 0.02 | -0.46 | -0.40 |
| Age: 50-64 | 0.43 | 0.01 | 0.41 | 0.46 |
| Age: 65+ | 1.07 | 0.02 | 1.04 | 1.10 |
| Acute Kidney Injury | -0.23 | 0.03 | -0.29 | -0.17 |
| CCI: 1-2 | 0.06 | 0.01 | 0.03 | 0.09 |
| CCI: 3-4 | 0.13 | 0.02 | 0.08 | 0.18 |
| CCI: 5-10 | 0.17 | 0.03 | 0.11 | 0.23 |
| CCI: 11+ | 0.19 | 0.09 | 0.03 | 0.36 |
| Chronic Lung Disease | 0.04 | 0.02 | 0.00 | 0.07 |
| Congestive Heart Failure | 0.10 | 0.06 | -0.01 | 0.22 |
| COVID In Sep 2021 | 0.13 | 0.02 | 0.10 | 0.16 |
| COVID In Oct 2021 | 0.32 | 0.02 | 0.29 | 0.36 |
| COVID In Nov 2021 | 0.40 | 0.02 | 0.37 | 0.44 |
| COVID In Dec 2021 | 0.70 | 0.02 | 0.66 | 0.73 |
| COVID In Jan 2022 | 1.28 | 0.02 | 1.24 | 1.32 |
| Data Partner A | 0.35 | 0.03 | 0.29 | 0.40 |
| Data Partner B | 0.26 | 0.06 | 0.14 | 0.37 |
| Data Partner C | 0.35 | 0.04 | 0.27 | 0.43 |
| Data Partner D | 0.70 | 0.07 | 0.57 | 0.84 |
| Data Partner E | 0.46 | 0.06 | 0.36 | 0.57 |
| Data Partner F | 0.10 | 0.03 | 0.05 | 0.15 |
| Data Partner G | 0.72 | 0.03 | 0.66 | 0.79 |
| Data Partner H | -0.12 | 0.05 | -0.21 | -0.02 |
| Data Partner I | 0.54 | 0.05 | 0.45 | 0.64 |
| Data Partner K | 0.03 | 0.02 | -0.01 | 0.06 |
| Diabetes Complicated | 0.05 | 0.03 | 0.01 | 0.10 |
| Diabetes Uncomplicated | 0.08 | 0.02 | 0.04 | 0.12 |
| Gender: Male | -0.14 | 0.01 | -0.16 | -0.12 |
| Heart Failure | -0.09 | 0.05 | -0.19 | 0.01 |
| Immunocompromised | 0.57 | 0.05 | 0.47 | 0.68 |
| Intercept | -1.09 | 0.02 | -1.13 | -1.05 |
| Kidney Disease | 0.18 | 0.02 | 0.14 | 0.23 |
| Myocardial Infarction | -0.15 | 0.03 | -0.21 | -0.08 |
| Race/Ethnicity: Asian | 0.90 | 0.04 | 0.82 | 0.99 |
| Race/Ethnicity: Black | -0.43 | 0.02 | -0.47 | -0.40 |
| Race/Ethnicity: Hispanic | 0.14 | 0.02 | 0.10 | 0.18 |
| Race/Ethnicity: Pacific Islander | -0.20 | 0.14 | -0.47 | 0.06 |
| Race/Ethnicity: Other | -0.39 | 0.04 | -0.46 | -0.32 |
| Race/Ethnicity: Unknown | -0.02 | 0.04 | -0.11 | 0.07 |
| SDOH: 0-45 | -0.30 | 0.07 | -0.44 | -0.16 |
| SDOH: 56-65 | 0.69 | 0.02 | 0.66 | 0.72 |
| SDOH: 65+ | 1.11 | 0.03 | 1.04 | 1.17 |
| SDOH: Missing | 0.20 | 0.03 | 0.15 | 0.25 |
| SDOH: Other | 0.20 | 0.02 | 0.17 | 0.24 |
| Tobacco Smoker | -0.55 | 0.03 | -0.60 | -0.50 |

**eTable 2. Clinic-Based IPTW Model Coefficients**

| Covariate | Coefficient | Standard Error | 95% CI Low | 95% CI High |
|---|---|---|---|---|
| Age: 18-24 | -0.75 | 0.04 | -0.83 | -0.67 |
| Age: 25-34 | -0.34 | 0.03 | -0.40 | -0.28 |
| Age: 50-64 | 0.24 | 0.03 | 0.18 | 0.29 |
| Age: 65+ | 0.69 | 0.03 | 0.63 | 0.76 |
| Acute Kidney Injury | -0.30 | 0.05 | -0.39 | -0.20 |
| CCI: 1-2 | 0.09 | 0.03 | 0.04 | 0.15 |
| CCI: 3-4 | 0.18 | 0.04 | 0.09 | 0.26 |
| CCI: 5-10 | 0.25 | 0.05 | 0.14 | 0.35 |
| CCI: 11+ | 0.28 | 0.12 | 0.04 | 0.51 |
| Chronic Lung Disease | 0.06 | 0.03 | 0.00 | 0.11 |
| Congestive Heart Failure | 0.10 | 0.10 | -0.10 | 0.30 |
| COVID In Sep 2021 | 0.17 | 0.04 | 0.10 | 0.24 |
| COVID In Oct 2021 | 0.33 | 0.04 | 0.25 | 0.42 |
| COVID In Nov 2021 | 0.38 | 0.05 | 0.29 | 0.47 |
| COVID In Dec 2021 | 0.89 | 0.04 | 0.82 | 0.96 |
| COVID In Jan 2022 | 1.29 | 0.03 | 1.23 | 1.35 |
| Data Partner A | 0.28 | 0.03 | 0.22 | 0.33 |
| Data Partner C | 0.28 | 0.04 | 0.20 | 0.36 |
| Data Partner D | 0.62 | 0.07 | 0.49 | 0.76 |
| Data Partner G | 0.62 | 0.03 | 0.55 | 0.69 |
| Data Partner I | 0.59 | 0.05 | 0.49 | 0.69 |
| Diabetes Complicated | 0.10 | 0.05 | 0.01 | 0.19 |
| Diabetes Uncomplicated | 0.09 | 0.04 | 0.01 | 0.17 |
| Gender: Male | -0.19 | 0.02 | -0.23 | -0.14 |
| Heart Failure | -0.09 | 0.09 | -0.27 | 0.09 |
| Immunocompromised | 0.81 | 0.08 | 0.67 | 0.96 |
| Intercept | -0.95 | 0.04 | -1.03 | -0.88 |
| Kidney Disease | 0.19 | 0.04 | 0.11 | 0.27 |
| Myocardial Infarction | -0.11 | 0.05 | -0.22 | -0.01 |
| Race/Ethnicity: Asian | 0.66 | 0.09 | 0.49 | 0.83 |
| Race/Ethnicity: Black | -0.36 | 0.03 | -0.41 | -0.31 |
| Race/Ethnicity: Hispanic | -0.24 | 0.04 | -0.33 | -0.16 |
| Race/Ethnicity: Pacific Islander | -0.52 | 0.25 | -1.02 | -0.02 |
| Race/Ethnicity: Other | -0.68 | 0.07 | -0.82 | -0.55 |
| Race/Ethnicity: Unknown | -0.23 | 0.07 | -0.36 | -0.10 |
| SDOH: 0-45 | -0.51 | 0.18 | -0.87 | -0.16 |
| SDOH: 56-65 | 0.69 | 0.02 | 0.65 | 0.74 |
| SDOH: 65+ | 1.33 | 0.06 | 1.22 | 1.44 |
| SDOH: Missing | -0.40 | 0.08 | -0.56 | -0.25 |
| SDOH: Other | -0.06 | 0.05 | -0.15 | 0.04 |
| Tobacco Smoker | -0.65 | 0.03 | -0.72 | -0.58 |

**eTable 3. Model-based Logistic Regression, All Coefficients**

| Covariate | Coefficient | Standard Error | 95% CI Low | 95% CI High |
|---|---|---|---|---|
| Age: 18-24 | -1.45 | 0.14 | -1.72 | -1.17 |
| Age: 25-34 | -0.80 | 0.07 | -0.94 | -0.65 |
| Age: 50-64 | 0.17 | 0.05 | 0.08 | 0.27 |
| Age: 65+ | 0.16 | 0.06 | 0.04 | 0.28 |
| Acute Kidney Injury | 0.27 | 0.09 | 0.09 | 0.46 |
| Chronic Lung Disease | 1.06 | 0.05 | 0.96 | 1.15 |
| Congestive Heart Failure | -0.25 | 0.16 | -0.56 | 0.06 |
| COVID In Sep 2021 | -0.06 | 0.05 | -0.16 | 0.05 |
| COVID In Oct 2021 | -0.21 | 0.07 | -0.34 | -0.07 |
| COVID In Nov 2021 | -0.17 | 0.07 | -0.32 | -0.03 |
| COVID In Dec 2021 | -0.32 | 0.07 | -0.45 | -0.18 |
| COVID In Jan 2022 | -0.78 | 0.07 | -0.92 | -0.64 |
| Data Partner A | 0.11 | 0.08 | -0.05 | 0.27 |
| Data Partner B | -0.18 | 0.22 | -0.60 | 0.24 |
| Data Partner C | -0.12 | 0.13 | -0.37 | 0.13 |
| Data Partner D | -0.78 | 0.23 | -1.22 | -0.33 |
| Data Partner E | -0.61 | 0.23 | -1.06 | -0.17 |
| Data Partner F | -0.50 | 0.07 | -0.64 | -0.36 |
| Data Partner G | -0.02 | 0.12 | -0.26 | 0.21 |
| Data Partner H | 0.01 | 0.13 | -0.26 | 0.27 |
| Data Partner I | 0.94 | 0.14 | 0.67 | 1.21 |
| Data Partner K | -1.54 | 0.05 | -1.64 | -1.43 |
| Diabetes Complicated | -0.08 | 0.09 | -0.26 | 0.10 |
| Diabetes Uncomplicated | 0.28 | 0.08 | 0.13 | 0.43 |
| Gender: Male | -0.21 | 0.04 | -0.30 | -0.13 |
| Heart Failure | 0.64 | 0.13 | 0.38 | 0.90 |
| Immunocompromised | 0.00 | 0.12 | -0.23 | 0.23 |
| Intercept | -3.08 | 0.07 | -3.21 | -2.95 |
| Kidney Disease | 0.03 | 0.09 | -0.16 | 0.21 |
| Myocardial Infarction | -0.06 | 0.10 | -0.26 | 0.13 |
| Race/Ethnicity: Asian | 0.11 | 0.27 | -0.41 | 0.64 |
| Race/Ethnicity: Black | 0.01 | 0.05 | -0.10 | 0.11 |
| Race/Ethnicity: Hispanic | -0.14 | 0.09 | -0.31 | 0.03 |
| Race/Ethnicity: Pacific Islander | 0.38 | 0.28 | -0.16 | 0.93 |
| Race/Ethnicity: Other | -0.43 | 0.17 | -0.77 | -0.09 |
| Race/Ethnicity: Unknown | -0.12 | 0.17 | -0.45 | 0.20 |
| Tobacco Smoker | -0.03 | 0.07 | -0.16 | 0.10 |
| Vaccination Status | -0.36 | 0.04 | -0.44 | -0.28 |

**eTable 4. Clinic-based Logistic Regression, All Coefficients**

| Covariate | Coefficient | Standard Error | 95% CI Low | 95% CI High |
|---|---|---|---|---|
| Age: 18-24 | -1.19 | 0.25 | -1.67 | -0.71 |
| Age: 25-34 | -0.63 | 0.15 | -0.93 | -0.33 |
| Age: 50-64 | 0.16 | 0.12 | -0.06 | 0.39 |
| Age: 65+ | 0.25 | 0.13 | 0.00 | 0.50 |
| Acute Kidney Injury | 0.31 | 0.19 | -0.06 | 0.68 |
| Chronic Lung Disease | 0.69 | 0.11 | 0.48 | 0.89 |
| Congestive Heart Failure | 0.34 | 0.36 | -0.35 | 1.04 |
| COVID In Sep 2021 | -0.26 | 0.12 | -0.49 | -0.02 |
| COVID In Oct 2021 | -0.41 | 0.17 | -0.75 | -0.08 |
| COVID In Nov 2021 | -0.25 | 0.16 | -0.57 | 0.06 |
| COVID In Dec 2021 | -0.54 | 0.15 | -0.83 | -0.24 |
| COVID In Jan 2022 | -0.89 | 0.13 | -1.15 | -0.64 |
| Data Partner A | -0.17 | 0.13 | -0.43 | 0.09 |
| Data Partner C | 0.19 | 0.18 | -0.16 | 0.54 |
| Data Partner D | -0.06 | 0.27 | -0.59 | 0.46 |
| Data Partner G | 0.33 | 0.13 | 0.08 | 0.59 |
| Data Partner I | 1.34 | 0.16 | 1.03 | 1.65 |
| Diabetes Complicated | -0.10 | 0.19 | -0.48 | 0.28 |
| Diabetes Uncomplicated | 0.11 | 0.18 | -0.24 | 0.46 |
| Gender: Male | -0.11 | 0.10 | -0.30 | 0.08 |
| Heart Failure | -0.23 | 0.34 | -0.90 | 0.44 |
| Immunocompromised | 0.06 | 0.24 | -0.42 | 0.53 |
| Intercept | -3.73 | 0.12 | -3.97 | -3.50 |
| Kidney Disease | 0.16 | 0.15 | -0.14 | 0.47 |
| Myocardial Infarction | -0.48 | 0.24 | -0.96 | -0.01 |
| Race/Ethnicity: Asian | 0.32 | 0.31 | -0.28 | 0.92 |
| Race/Ethnicity: Black | -0.40 | 0.13 | -0.67 | -0.14 |
| Race/Ethnicity: Hispanic | -0.11 | 0.19 | -0.47 | 0.26 |
| Race/Ethnicity: Other | 0.03 | 0.25 | -0.47 | 0.53 |
| Race/Ethnicity: Unknown | 0.06 | 0.25 | -0.43 | 0.55 |
| Tobacco Smoker | -0.22 | 0.15 | -0.51 | 0.08 |
| Vaccination Status | -0.36 | 0.09 | -0.53 | -0.20 |

**eTable 5. Model-based Proportional Hazards, All Coefficients**

| Covariate | Coefficient | Standard Error | 95% CI Low | 95% CI High |
|---|---|---|---|---|
| Age: 18-24 | -2.70 | 0.42 | -3.52 | -1.88 |
| Age: 25-34 | -0.89 | 0.10 | -1.08 | -0.70 |
| Age: 50-64 | 0.25 | 0.07 | 0.13 | 0.38 |
| Age: 65+ | 0.27 | 0.08 | 0.12 | 0.42 |
| Acute Kidney Injury | 0.18 | 0.12 | -0.05 | 0.41 |
| Chronic Lung Disease | 1.02 | 0.06 | 0.90 | 1.14 |
| Congestive Heart Failure | -0.28 | 0.16 | -0.59 | 0.04 |
| COVID In Sep 2021 | -0.06 | 0.07 | -0.21 | 0.08 |
| COVID In Oct 2021 | -0.11 | 0.09 | -0.30 | 0.07 |
| COVID In Nov 2021 | 0.09 | 0.09 | -0.09 | 0.27 |
| COVID In Dec 2021 | 0.16 | 0.09 | -0.03 | 0.34 |
| COVID In Jan 2022 | -0.19 | 0.11 | -0.42 | 0.03 |
| Data Partner A | 0.33 | 0.09 | 0.15 | 0.51 |
| Data Partner B | -0.32 | 0.23 | -0.77 | 0.12 |
| Data Partner C | -0.13 | 0.16 | -0.44 | 0.18 |
| Data Partner D | 1.46 | 0.82 | -0.15 | 3.06 |
| Data Partner E | -0.52 | 0.38 | -1.28 | 0.23 |
| Data Partner F | -1.10 | 0.18 | -1.45 | -0.74 |
| Data Partner G | -0.10 | 0.18 | -0.46 | 0.26 |
| Data Partner H | 0.21 | 0.15 | -0.08 | 0.51 |
| Data Partner I | 0.64 | 0.20 | 0.25 | 1.03 |
| Data Partner K | -1.13 | 0.07 | -1.27 | -0.98 |
| Diabetes Complicated | -0.04 | 0.10 | -0.24 | 0.17 |
| Diabetes Uncomplicated | 0.27 | 0.09 | 0.10 | 0.44 |
| Gender: Male | 0.24 | 0.11 | 0.02 | 0.46 |
| Heart Failure | 0.65 | 0.15 | 0.34 | 0.95 |
| Immunocompromised | 0.15 | 0.16 | -0.16 | 0.47 |
| Kidney Disease | -0.07 | 0.11 | -0.28 | 0.14 |
| Myocardial Infarction | 0.03 | 0.12 | -0.19 | 0.26 |
| Race/Ethnicity: Asian | 0.33 | 0.38 | -0.41 | 1.07 |
| Race/Ethnicity: Black | 0.04 | 0.07 | -0.09 | 0.17 |
| Race/Ethnicity: Hispanic | -0.12 | 0.12 | -0.35 | 0.11 |
| Race/Ethnicity: Other | -0.31 | 0.20 | -0.71 | 0.09 |
| Race/Ethnicity: Unknown | -0.16 | 0.27 | -0.69 | 0.37 |
| Tobacco Smoker | 0.02 | 0.09 | -0.16 | 0.19 |
| Vaccination Status | -0.47 | 0.05 | -0.58 | -0.37 |
| Time * Gender: Male | -0.00 | 0.00 | -0.01 | -0.00 |
| Time * Age: 18-24 | 0.01 | 0.00 | 0.00 | 0.02 |
| Time * Data Partner D | -0.02 | 0.01 | -0.04 | -0.00 |
| Time * Data Partner F | 0.01 | 0.00 | 0.00 | 0.01 |

**eTable 6. Clinic-based Proportional Hazards, All Coefficients**

| Covariate | Coefficient | Standard Error | 95% CI Low | 95% CI High |
|---|---|---|---|---|
| Age: 18-24 | -1.26 | 0.30 | -1.86 | -0.67 |
| Age: 25-34 | -0.55 | 0.18 | -0.91 | -0.19 |
| Age: 50-64 | 0.26 | 0.14 | -0.01 | 0.53 |
| Age: 65+ | 0.31 | 0.16 | 0.00 | 0.62 |
| Acute Kidney Injury | 0.41 | 0.19 | 0.03 | 0.78 |
| Chronic Lung Disease | 0.76 | 0.12 | 0.52 | 0.99 |
| Congestive Heart Failure | 0.09 | 0.39 | -0.68 | 0.86 |
| COVID In Sep 2021 | -0.06 | 0.14 | -0.34 | 0.21 |
| COVID In Oct 2021 | -0.27 | 0.20 | -0.67 | 0.13 |
| COVID In Nov 2021 | -0.13 | 0.21 | -0.55 | 0.29 |
| COVID In Dec 2021 | -0.29 | 0.20 | -0.67 | 0.09 |
| COVID In Jan 2022 | -0.48 | 0.15 | -0.76 | -0.19 |
| Data Partner A | -0.03 | 0.14 | -0.30 | 0.23 |
| Data Partner C | 0.26 | 0.22 | -0.18 | 0.69 |
| Data Partner D | -0.09 | 0.29 | -0.66 | 0.48 |
| Data Partner G | 0.18 | 0.17 | -0.14 | 0.51 |
| Data Partner I | 1.32 | 0.17 | 0.98 | 1.66 |
| Diabetes Complicated | -0.10 | 0.23 | -0.56 | 0.35 |
| Diabetes Uncomplicated | 0.10 | 0.21 | -0.31 | 0.51 |
| Gender: Male | -0.12 | 0.10 | -0.32 | 0.09 |
| Heart Failure | 0.00 | 0.36 | -0.71 | 0.71 |
| Immunocompromised | 0.09 | 0.24 | -0.39 | 0.56 |
| Kidney Disease | 0.16 | 0.16 | -0.16 | 0.47 |
| Myocardial Infarction | -0.56 | 0.25 | -1.04 | -0.08 |
| Race/Ethnicity: Asian | 0.52 | 0.44 | -0.34 | 1.38 |
| Race/Ethnicity: Black | -0.38 | 0.15 | -0.68 | -0.09 |
| Race/Ethnicity: Hispanic | -0.16 | 0.22 | -0.58 | 0.27 |
| Race/Ethnicity: Other | 0.13 | 0.30 | -0.45 | 0.72 |
| Race/Ethnicity: Unknown | 0.23 | 0.31 | -0.38 | 0.85 |
| Tobacco Smoker | -0.26 | 0.18 | -0.61 | 0.09 |
| Vaccination Status | -0.41 | 0.10 | -0.60 | -0.22 |

**eFigure 1. Differences in standardized covariates in the model-based cohort between vaccinated and unvaccinated groups before (unadjusted) and after (adjusted) IPTW.**

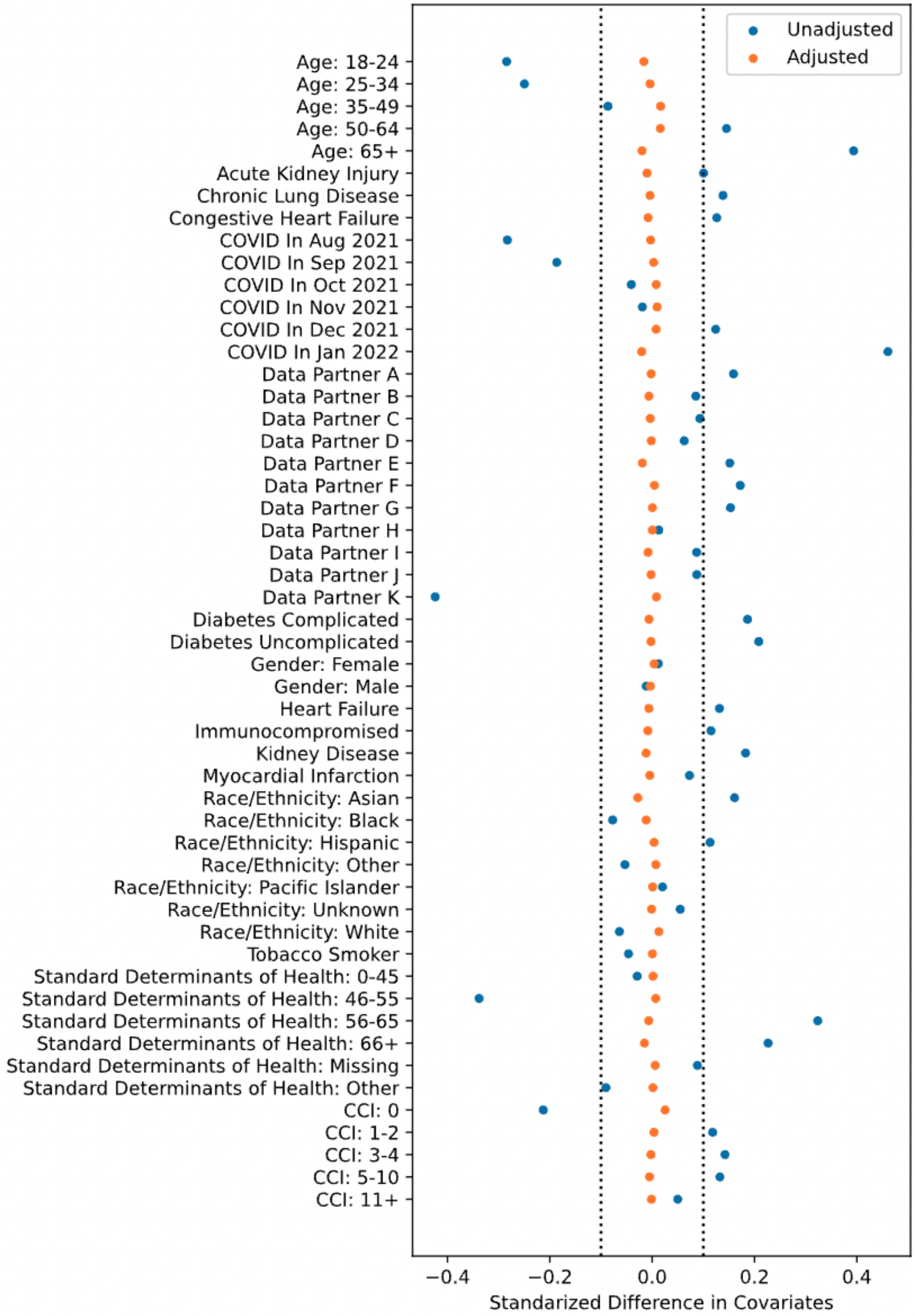

**eFigure 2. Differences in standardized covariates in the clinic-based cohort between vaccinated and unvaccinated groups before (unadjusted) and after (adjusted) IPTW.**

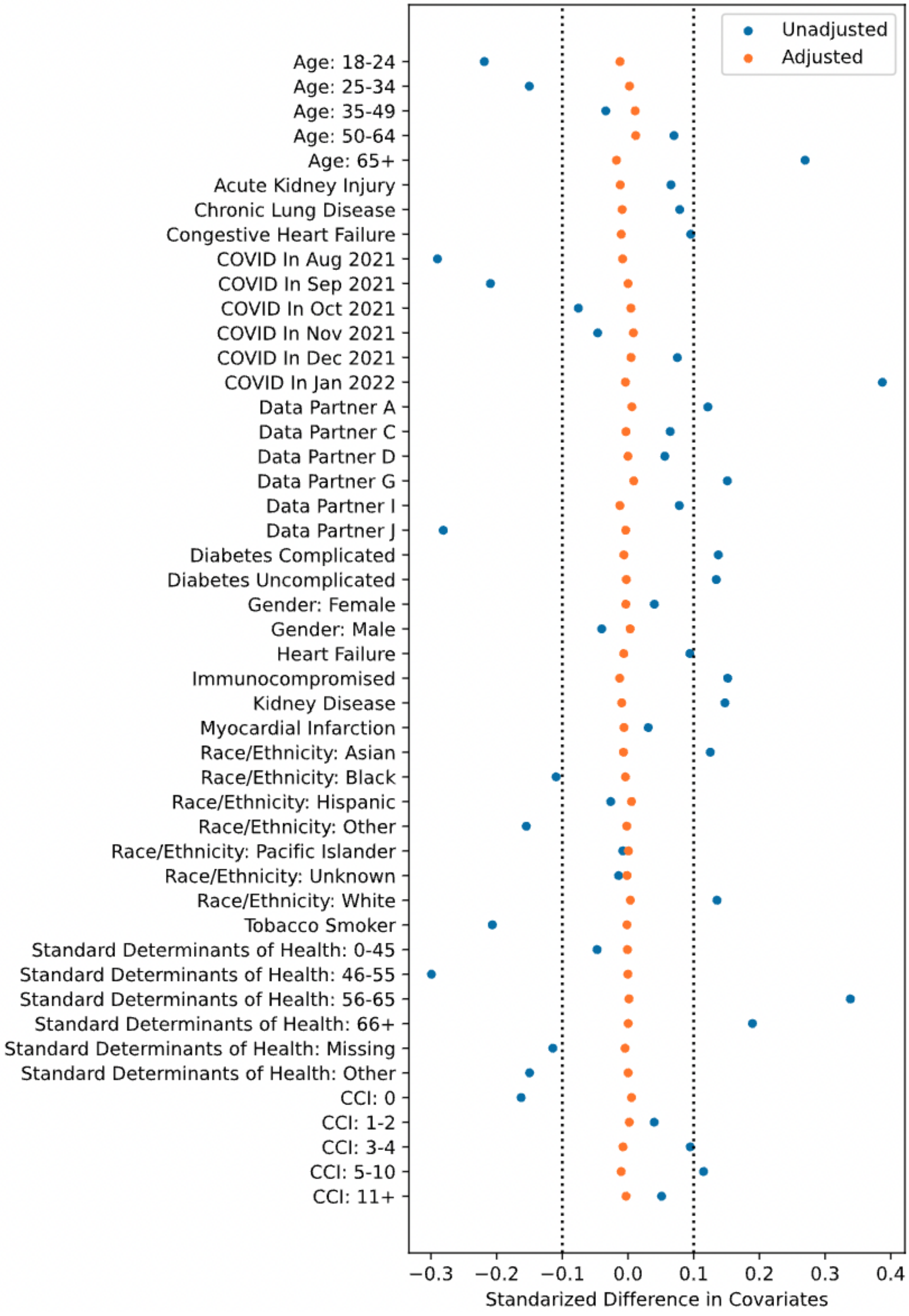

**eTable 7. Associations of Age with Vaccination and Long COVID**

| | Age at COVID Index Date | Overall | Fully Vaccinated | Unvaccinated | With Long COVID | Without Long COVID |
|---|---|---|---|---|---|---|
| Model-based Cohort | All | 199498 (100.0) | 87099 (100.0) | 112399 (100.0) | 3405 (100.0) | 196093 (100.0) |
| | 18-24 | 20701 (10.4) | 4922 (5.7) | 15779 (14.0) | 71 (2.1) | 20630 (10.5) |
| | 25-34 | 36729 (18.4) | 11382 (13.1) | 25347 (22.6) | 292 (8.6) | 36437 (18.6) |
| | 35-49 | 53883 (27.0) | 21646 (24.9) | 32237 (28.7) | 984 (28.9) | 52899 (27.0) |
| | 50-64 | 50887 (25.5) | 25332 (29.1) | 25555 (22.7) | 1175 (34.5) | 49712 (25.4) |
| | 65+ | 37298 (18.7) | 23817 (27.3) | 13481 (12.0) | 883 (25.9) | 36415 (18.6) |
| Clinic-based Cohort | All | 47752 (100.0) | 26567 (100.0) | 21185 (100.0) | 706 (100.0) | 47046 (100.0) |
| | 18-24 | 4522 (9.5) | 1753 (6.6) | 2769 (13.1) | 22 (3.1) | 4500 (9.6) |
| | 25-34 | 8557 (17.9) | 4078 (15.3) | 4479 (21.1) | 68 (9.6) | 8489 (18.0) |
| | 35-49 | 12445 (26.1) | 6748 (25.4) | 5697 (26.9) | 194 (27.5) | 12251 (26.0) |
| | 50-64 | 12356 (25.9) | 7235 (27.2) | 5121 (24.2) | 232 (32.9) | 12124 (25.8) |
| | 65+ | 9872 (20.7) | 6753 (25.4) | 3119 (14.7) | 190 (26.9) | 9682 (20.6) |

**eFigure 3. Sensitivity analysis results for logistic regression (LR) and proportional hazards (PH), full LC Model threshold range.** Odds ratios for the vaccination coefficient are shown for LR, hazard ratios for the vaccination coefficient are shown for PH. Dashed lines are results from the clinic-based cohort, where the LC model cutoff is not relevant. Solid lines are results from the model-based cohort.

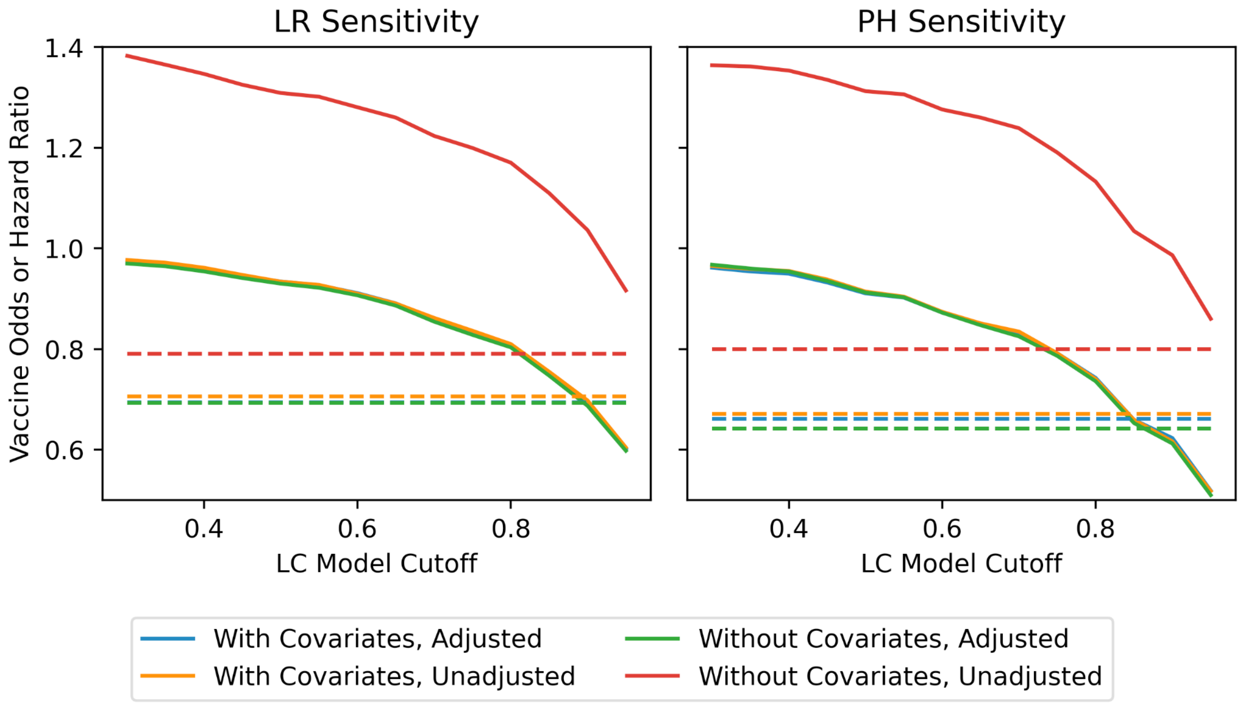

### eDiscussion

We do not interpret the associations between vaccination and long COVID here as causal, as we fail to fully account for two important conditions: unconfoundedness and latent variables.

#### Unconfoundedness

Under certain assumptions, associations in IPTW-adjusted models can be interpreted as causal effects, even when treatment is not randomly assigned.[1,2] If we are willing to assume that there are no unmeasured confounders, IPTW satisfies the condition of exchangeability: the treatment and control groups differ in outcome only due to the treatment. We attempt to satisfy this assumption by controlling for the measured confounders outlined in eTables 1 and 2 in our treatment model. Nevertheless, we do not assume that there are no further confounders, largely due to the field's nascent understanding of long COVID. For example, a small cohort study suggested that reactivation of latent viruses may contribute to long COVID, but we do not include past viral infections in the treatment model.[3] Furthermore, some of the latent variables in our causal model are unobserved confounders, as discussed further below.

#### Latent Variables

To assess the feasibility of interpreting our results causally, we developed a simplified, theoretical causal model of COVID-19 and long COVID. This model is illustrated with a directed acyclic graph (DAG) in eFigure 4 and reveals latent variables that present an obstacle to estimating the causal effect of COVID-19 vaccination on long COVID. Of particular concern is patient propensity to seek healthcare, which affects the likelihood that both COVID-19 vaccination and long COVID will be observed, and is common in research using electronic health records.[4]

**eFigure 4: Directed acyclic graph (DAG) of a simplified, theoretical causal model of COVID-19 vaccination and long COVID.**

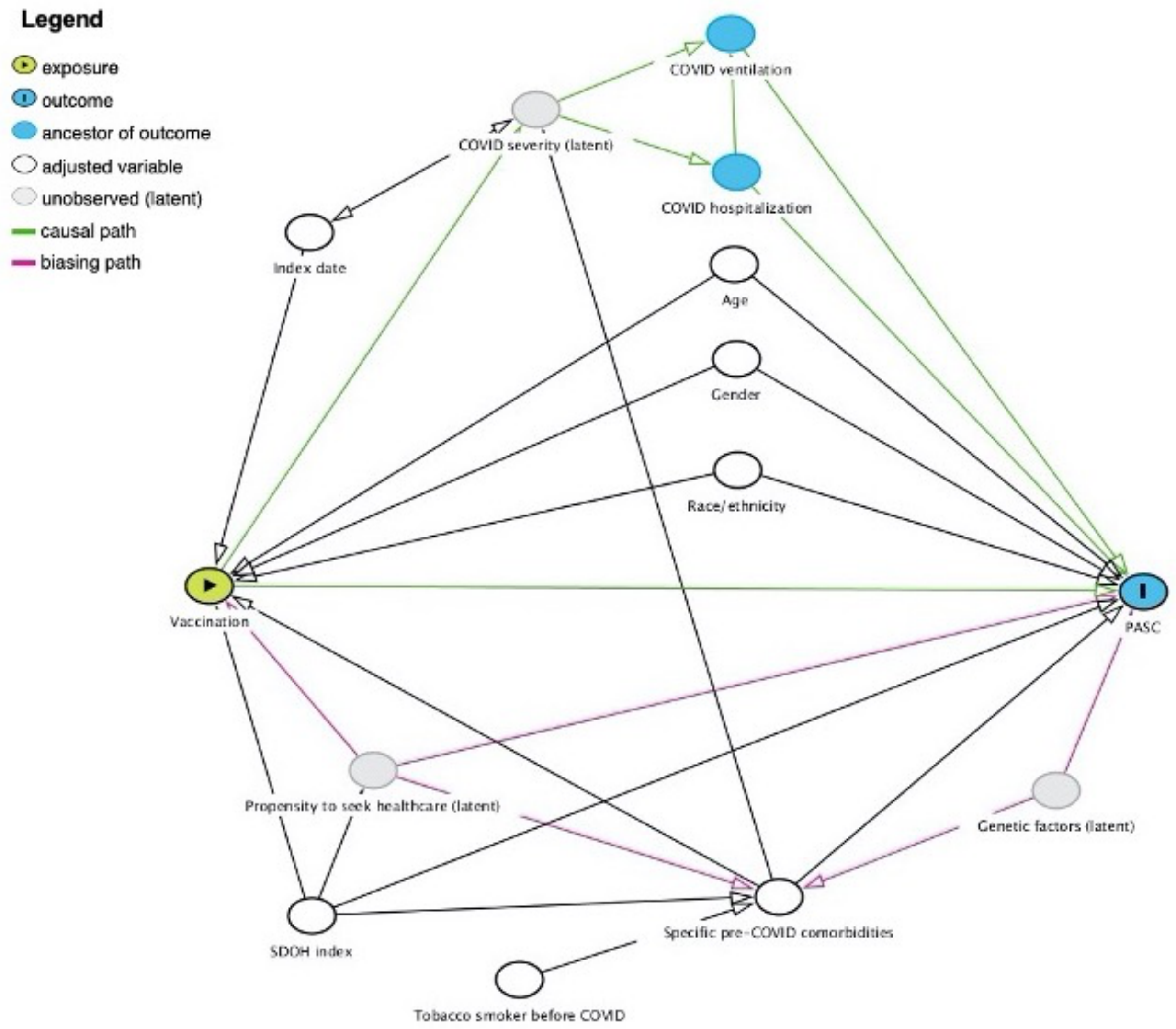
